# Lessons learned from Vietnam’s COVID-19 response: the role of adaptive behavior change and testing in epidemic control

**DOI:** 10.1101/2020.12.18.20248454

**Authors:** Quang D. Pham, Robyn M. Stuart, Thuong V. Nguyen, Quang C. Luong, Dai Q. Tran, Thai Q. Pham, Lan T. Phan, Tan Q. Dang, Duong N. Tran, Hung T. Do, Dina Mistry, Daniel J. Klein, Romesh G. Abeysuriya, Assaf P. Oron, Cliff C. Kerr

## Abstract

**Background:** Vietnam has emerged as one of the world’s leading success stories in responding to COVID-19. After prolonged zero-low transmission, a summer outbreak of unknown source at Da Nang caused the country’s first COVID-19 deaths, but was quickly suppressed. Vietnam recently reopened its borders to international travelers. Understanding the attendant risks and how to minimize them is crucial as Vietnam moves into this new phase.

**Methods:** We create an agent-based model of COVID-19 in Vietnam, using regional testing data and a detailed linelist of the 1,014 COVID-19 cases, including 35 deaths, identified across Vietnam. We investigate the Da Nang outbreak, and quantify the risk of another outbreak under different assumptions about behavioral/policy responses and ongoing testing.

**Results:** The Da Nang outbreak, although rapidly contained once detected, nevertheless caused significant community transmission before it was detected; higher symptomatic testing could have mitigated this. If testing levels do not increase, the adoption of past policies in response to newly-detected cases may reduce the size of potential outbreaks but will not prevent them. Compared to a baseline symptomatic testing rate of 10%, we estimate half as many infections under a 20% testing rate, and a quarter as many with 40-50% testing rates, over the four months following border reopenings.

**Conclusions:** Vietnam’s success in controlling COVID-19 is largely attributable to its rapid response to detected outbreaks, but the speed of response could be improved even further with higher levels of symptomatic testing.

## Introduction

With its first confirmed case on January 23, 2020, Vietnam was among the first ten countries affected by the worldwide COVID-19 pandemic.^1^ Two weeks later, only 150 cases had been reported outside of mainland China, but 10 (7%) of these were in Vietnam, placing it in the top ten most affected countries.^2^ However, one year later the situation was very different: with just 1300 cumulative cases and 0.4 deaths per million inhabitants, Vietnam ranked among the five countries with the lowest COVID-19 disease burden, and among the three countries with lowest overall mortality.^3^ Understanding how transmission has been kept so low and whether it can be maintained as such is crucial, especially as Vietnam reopens its borders to international arrivals.

Vietnam has been widely praised for this notable success in suppressing the spread of COVID-19.^4,5^ The initial response to COVID-19 was characterized by a series of measures to prevent onward transmission following importation (Figure 1). The government closed its border with China and suspended air travel from China on January 28, 2020. The travel restriction policy extended to other COVID-19 affected countries in mid-February, and all international travel by commercial airlines was suspended from March 25 onwards. Additional border control measures included screening at border gates and quarantine of all travelers entering Vietnam. The country closed all land border crossings and transportation lines with the three neighboring countries of China, Laos, and Cambodia, canceled train services with China, and denied permission to dock cruise ships. Schools and universities closed initially for the 1-week Tet holidays (the Vietnamese Lunar New Year), and the closure was extended in response to the COVID-19 epidemic. In March, the government issued a mask-wearing mandate, a ban on public gatherings (including entertainment, cultural/sporting events and non-essential businesses), and a 2-meter physical distancing recommendation.

**Figure 1.**
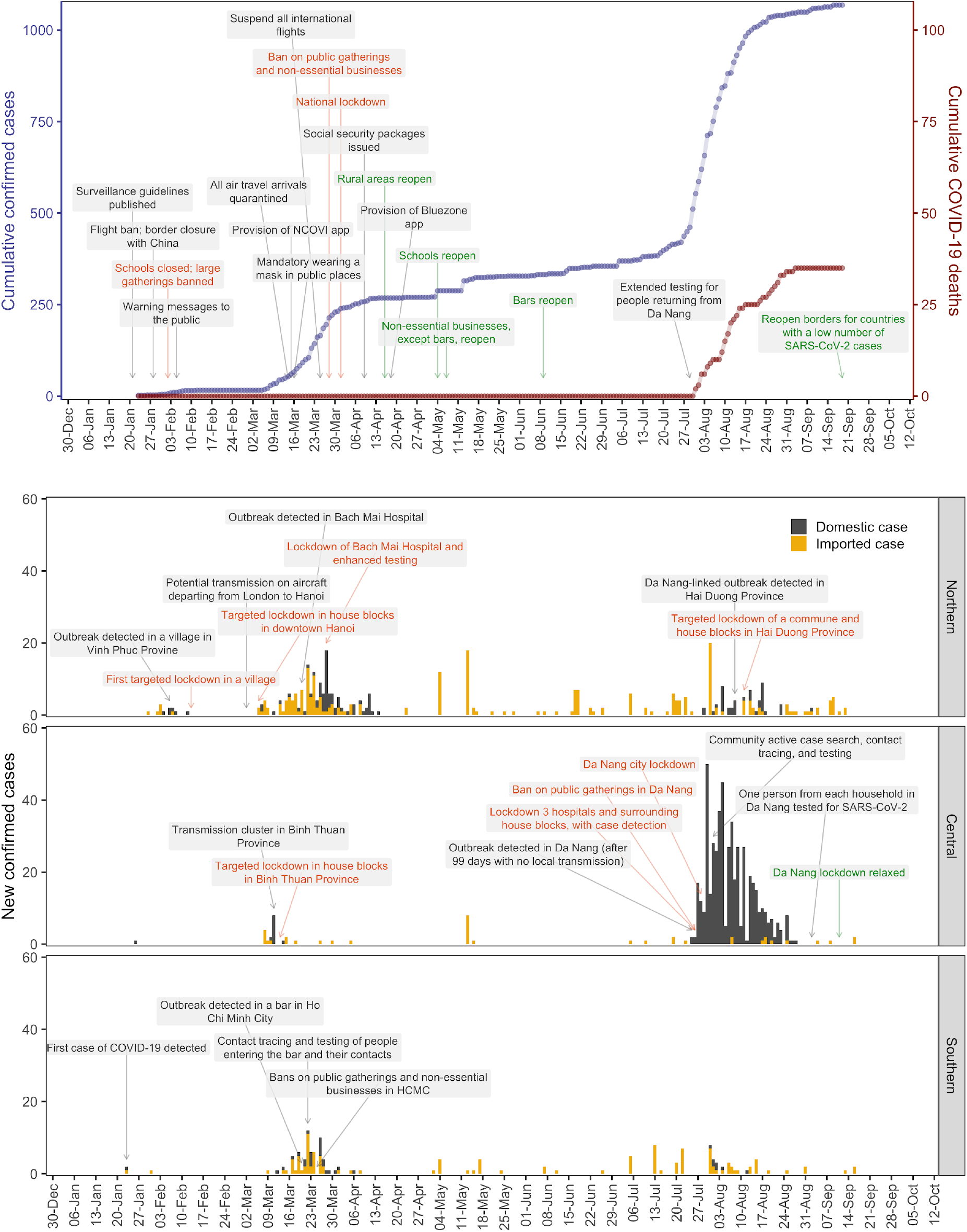
(A) Cumulative known COVID-19 cases (blue) and deaths (maroon) in Vietnam, January–September 2020. Key national pandemic response measures are shown, with the text centered on the date they were enacted. Measures marked in red and green indicate lockdowns and their relaxation, respectively. (B) Regional epidemiological curves in Vietnam with case source (domestic or imported) indicated, January–September 2020. Key local and regional response measures are shown, color-coded as in Figure 1a.

Together with enhancement of communication to the public, surveillance, isolation and quarantine, testing, and contacting tracing, Vietnam adopted targeted lockdowns early in the pandemic. After several separate epidemics were detected in different locations over a short period of time, the government imposed a national lockdown from April 1 to April 15 to curb transmission of COVID-19 in the community. These measures helped Vietnam to stay free of community transmission of COVID-19 for 99 days. On July 25, a cluster of domestic cases was detected at a provincial hospital in Da Nang in Central Vietnam, a popular domestic tourism destination. The Da Nang region was locked down on July 28, but the outbreak still spread rapidly around the city, with cases also showing up in 14 other provinces across Vietnam through various forms of transportation, including airplanes, trains and buses, during the peak tourist season in the city. By late August the Da Nang outbreak had been suppressed, but not before causing over 500 known cases, more than Vietnam’s entire case count through July 24, as well as the country’s first recorded COVID-19 deaths. In view of the Da Nang experience and the continued failure of many countries to curb the pandemic, Vietnam has been reluctant to reopen its borders for international travelers.

Mathematical modelling has formed a crucial element of the global response to the COVID-19 pandemic since its onset.^6^ Vietnam’s exemplary COVID-19 response, informed by its previous experience with SARS, has meant that thus far it has not relied upon modelling as much as countries suffering higher pandemic burden. However, Vietnam’s relatively unique COVID-19 situation means that the problem of reopening borders with zero local transmission and robust pandemic response systems has not received much modelling attention. The available literature on the risks associated with international travel has determined that border closures were an important part of limiting the spread of the virus,^7,8^ but only a small number of other studies have examined the risks associated with reopening borders,^9,10^ and only one has examined the impact of different quarantine policies, finding that quarantine periods of 8 or 14 days followed by a negative test result could reduce the number of infectious arrivals released into the community by a median 94% or 99% respectively compared to a no quarantine, no test scenario.^11^

To address Vietnam’s needs as it reopens its borders, we have set out to model adaptive behavioral changes in response to new information about the state of the epidemic. On July 25, Vietnam had recorded 99 days with no locally transmitted cases. A survey of ∼95,000 people conducted the following day indicated that the proportion of people who reported frequently and sometimes using a face mask over the preceding 2 weeks were 35% and 29%.^12^ However, over the following week in response to reports of the Da Nang outbreak, public behavior changed rapidly: by the time the next survey was conducted on August 2, the proportions of people who reported frequently and sometimes using a face mask over the preceding 2 weeks were 90% and 5% respectively.^13^ Feedback mechanisms between the number of reported cases and people’s behavior has rarely been incorporated directly into modelling frameworks, although some studies have included dynamic interventions based on trigger thresholds.^14–16^ Including this feature for Vietnam is essential in order to be able to capture the evolution of the Da Nang outbreak, and to make reasonable estimates of the probability of future outbreaks in Vietnam and countries with similar COVID-19 performance (e.g., Taiwan and New Zealand) following the reopening of borders.

In this paper, we use detailed data from the outbreak of COVID-19 in the province of Da Nang in August of 2020 to calibrate a stochastic agent-based model of COVID-19 transmission for Vietnam. We use this model in a framework for evaluating the risk of epidemic rebound in Vietnam following the reopening of international borders.

## Methods

### Data inputs

Testing and patient data were gathered from the National Institute of Hygiene and Epidemiology, an organization that supports public health activities in Vietnam, and from the General Department of Preventive Medicine (GDPM), a Ministry of Health affiliated department, respectively. The daily and cumulative numbers of tests and diagnosed cases separated by geographic regions of laboratories, including the North, Central, and South, were collected via a national reporting system of COVID-19 testing. For each patient diagnosed, information was recorded on age, gender, nationality, geographical origin, case classification (imported or domestic cases), date of diagnosis, signs and symptoms they experienced, date of illness onset, date of isolation, date of hospitalization, the number of patient’s close contacts getting infected with SARS-CoV-2, any complications developed during hospital administration, and date of death if it occurred. In total, we obtained a detailed line-list of 1,014 de-duplicated laboratory-confirmed COVID-19 patients, including 35 deaths, identified across Vietnam from January 23 to August 22.

In addition, data on incoming arrivals to airports in Vietnam’s South over the period from May 31, 2020 to October 24, 2020, were collected from the GDPM and the Pasteur Institute of Ho Chi Minh City, and analyzed for the purpose of quantifying the proportion of arrivals testing positive to COVID-19.

### Modelling the Da Nang outbreak

We use an open-source agent-based model called Covasim,^17^ documented in detail elsewhere,^18^ to create an agent-based model for Central Vietnam. We began by simulating a population representative of Central Vietnam by taking data on the age and sex composition of the population and using it to create a model population of agents with similar characteristics. Next, we used Covasim’s inbuilt methods to construct four distinct contact networks that assign these agents to households, schools, workplaces, and communities based on their ages.

Following the release of the initial lockdowns in April, Da Nang (a popular summer holiday destination) launched a campaign to attract domestic tourists, which led to a sharp increase in domestic tourist arrivals: ∼250,000 in May, 450,000 in June, ∼1.4 million in July = approx. 33,000/day over June 15 – July 25, 2020. Despite Vietnam’s aggressive response to the early waves of COVID-19, tests of 895 blood donors in Ho Chi Minh City in August showed low but non-zero prevalence of neutralizing antibodies against SARS-CoV-2 (2/895 ∼ 0.2%). We therefore initialized the model on June 15, 2020, and over the following 40 days until July 25, an average of 1 new infected person per day (or 0.003% of incoming tourists; drawn from a negative binomial distribution with dispersion of 0.25) was introduced into the population. This low, steady stream of new cases reflects the influx of domestic tourists to Da Nang over this period, approximating the sudden concentration of a number of infected people into a single municipality.

To simulate the policy environment, we include parameter changes that capture Vietnam’s testing, tracing, isolation, quarantine, and lockdown strategies (summarized in Table S1). Over the period from June 15, 2020 to October 15, 2020, the model was calibrated to data from Central Vietnam on the number of tests, diagnosed cases, and deaths (as described in the section above). Calibration was performed by drawing 20 samples from a distribution of values for the per-contact transmission risk, running N=500 simulations for each to produce 10,000 trial simulations, and then retaining the 1% of these with the minimum absolute differences between the model projections and the data. Core parameter values and sources are documented in Table S1.

An important factor in modelling the Da Nang outbreak is the speed with which both official policy and the community reacted to the detection of new cases. Schools and workplaces in Da Nang were closed from 28 July (3 days after new cases were detected) until the 14th and 5th of September respectively, and a stay-at-home recommendation was issued (Table S1). In addition to survey data indicating that the proportion of people who reported frequently wearing a mask increased from 35% to 90% following the reported increase in locally transmitted cases, there was likely also an increase in vigilance with hygiene and distancing protocols. A case-control study on the use of masks and other personal protective measures in Thailand found that those who wore masks all the time were more likely to report that (a) their closest contacts were more than 1 meter away, (b) contact durations were limited to ≤15 minutes, and (c) they washed their hands often.^19^ Taking all these factors into account, the study found a negative association between the individual-level risk of COVID-19 transmission and wearing a mask all the time (adjusted odds ratio (aOR) 0.23, 95% CI 0.09–0.60), while wearing masks sometimes was not significantly associated with lower risk of infection (aOR 0.87, 95% CI 0.41–1.84). Using these estimates, we obtain a prior estimate for the overall individual-level impact of the reported increase in mask usage, corresponding to a reduction in the per-contact probability of transmission of 58% (26-73%) (details provided in Table S1). In the model, we assume that this applies to contacts that take place in the community, workplace, and school networks, but not in households as people are less likely to wear masks at home.

### Modelling the reopening of international borders

Firstly, to estimate the risk of an imported infection entering Vietnam and escaping quarantine, we analyze data on incoming arrivals to airports in Vietnam’s South and calculate the distribution of onward transmissions per infected arrival. These calculations take account of the quarantine protocols in place in Vietnam as of 20 November, 2020, which require all incoming international arrivals to Vietnam to: (a) test negative 3-5 days before departing; (b) disclose travel 14 days before departing; (c) complete a nasopharyngeal swab sample antigen test on arrival; (d) quarantine in a hotel for 7 days, after which they are tested for SARS-CoV-2 by RT-PCR or RT-LAMP; (e) quarantine at their residence for a further 7 days if both test results are negative; (f) continue to have their health monitored daily by a commune health worker over the remainder of their quarantine, with another nasopharyngeal swab sample collected at day 14 after arrival.^20^ This combined 14-day quarantine period is based on the distribution of the incubation period for COVID-19,^21–23^ and is consistent with current WHO recommendations.^24^ Vietnamese citizens who repatriated from foreign countries are quarantined for 14 days at government facilities and tested twice during the 14-day quarantine as the previous testing schedule.

Secondly, to model the risk of escaped cases causing an outbreak in Vietnam, we create a national model using the parameter values obtained via the calibration process for the Da Nang outbreak. Beginning from a point with no active cases in the community (which we take to be November 30, 2020, but which could theoretically be any point), we initialize 100 simulations and project forward by 4 months. On each day, the number of new imported cases on each day is drawn from a negative binomial distribution, with parameters based on the observed distribution of imported cases over the period from February 1–August 22, 2020, but scaled such that the mean number of imported cases is 10 times higher over the projection period when borders are open than when they were closed; the scale factor is based on the planned reopening schedule of Vietnam’s borders. For all future projections, we assume that schools and workplaces would be closed if more than 5 cases are detected. We also assume that all identified contacts of confirmed cases are tested regardless of symptoms; for those with COVID-19-like symptoms but no known history of contact with a case, we assume 10% will seek a test during periods of low transmission (based on an analysis of the testing data over February 1–August 22, 2020), but once more than 5 cases are detected, aggressive testing campaigns increase this to 90%.

The question of how policy and behavior will respond to increases in reported cases is highly relevant for analyzing the future trajectory of the epidemic in Vietnam. We therefore model three alternative scenarios:

1. Constant high compliance: for this scenario, we assume that the population will remain highly compliant with measures to stop transmission. We model this by assuming the reduction in transmission risk in response to the detection of new locally-transmitted cases in late July represented a permanent shift in behavior.
2. Increased complacency: for this scenario, we include another behavior change in the model, whereby the relative per-contact probability of transmission increases back to its pre-outbreak value after the 14-day average of new locally-transmitted cases falls below 2. Following the detection of new cases, the probability of someone with symptoms getting tested is assumed to increase as testing capacity is scaled up would result in policy actions comparable to those that have been implemented in the past (i.e., localized school and workplace closures following the detection of more than 5 locally-transmitted cases), but we assume that mask/NPI compliance would not increase again.
3. Self-regulating behavior: for this scenario, we allow the relative per-contact probability of transmission to be fully dynamic. We allow it to increase back to its pre-outbreak value whenever the 14-day average of new locally-transmitted cases falls below 2 (as in the “increased apathy” scenario above), but to decrease again if the daily number of new locally-transmitted cases increases above a certain threshold. This scenario is intended to capture behavior change in response to the information conveyed by case counts.

## Results

### Characteristics of the Da Nang outbreak

Figure 2 shows the model’s estimates of the characteristics of the outbreak in Central Vietnam, as well as comparisons to observed data where possible. According to our estimates, the outbreak in Central Vietnam had very likely begun well before the first cases were detected, with an estimated 1,480 (95% projected interval: 1,170-1,870) infections occurring between June 15 and July 25, 2020. Testing levels over this period were relatively low, averaging around 350 tests/day, but scaled up rapidly after cases were detected on July 25 to peak at around 17,000 tests/day by August 13, 2020. We estimate that the outbreak itself peaked with approximately 1,060 (95% projected interval: 890-1280) active infections on August 2, 2020.

**Figure 2.**
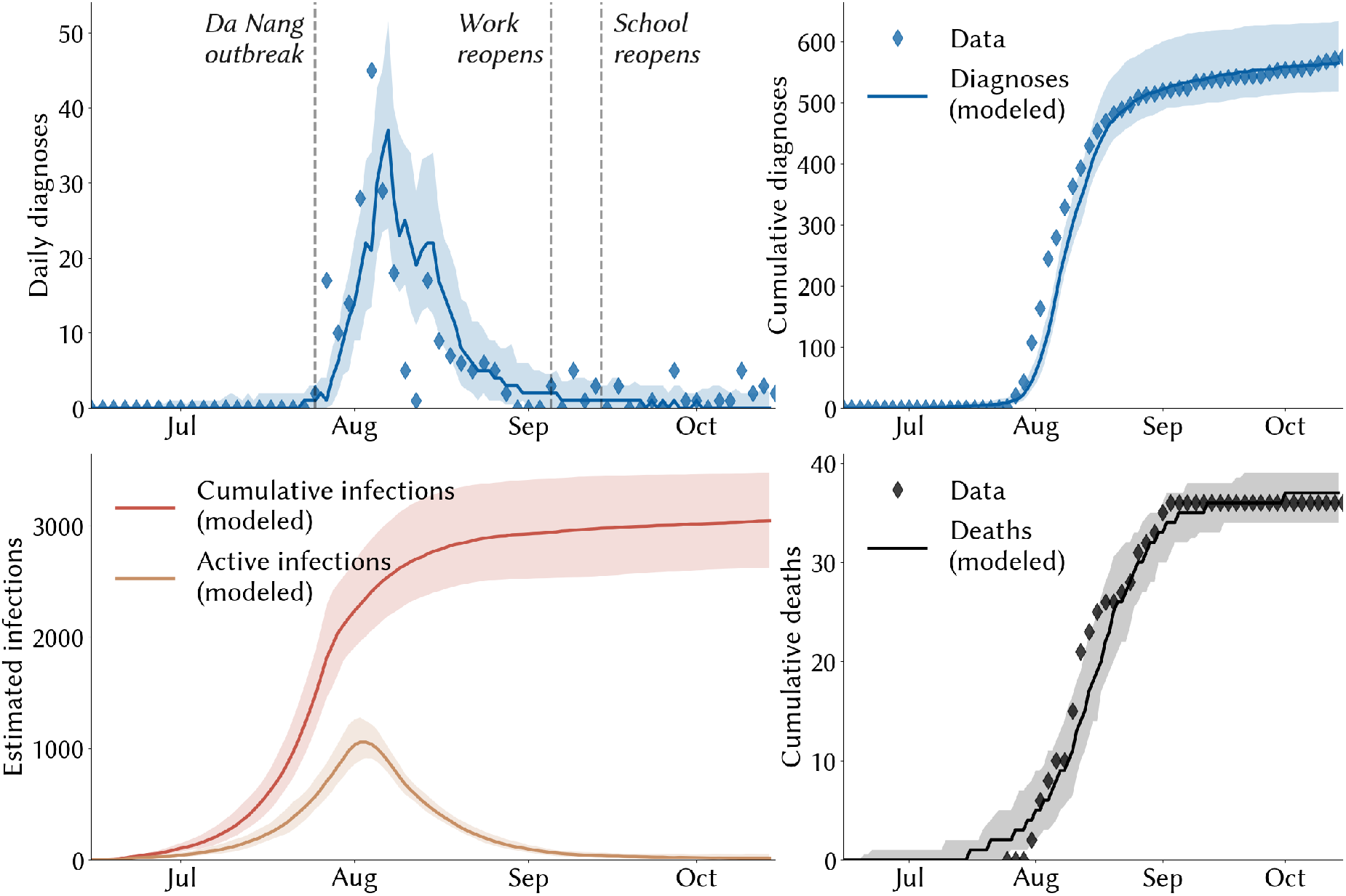
Fitting a microsimulation model to data on COVID-19 cases and deaths in Central Vietnam. Solid lines indicate the median model projections over 100 simulations; shaded areas indicate 95% projected intervals; diamonds indicate data. Until August 22, the daily diagnoses data includes local transmissions only, but after this point diagnoses could not be disaggregated, so data points on diagnoses after this date should be interpreted as overestimates of local transmission.

Over the entire outbreak, we estimate that around 3,020 (95% projected interval: 2,600-3,460) people were infected, of which only ∼20% were diagnosed. This relatively low case detection rate can largely be explained by the lower testing rates prior to July 25, 2020. Thereafter, we estimate that the majority of undiagnosed infections can be accounted for by asymptomatic transmission chains: almost half (47%, 95% projected interval: 42-51%) of undiagnosed infections that occurred after July 25 were asymptomatic, and of these, 72% (59-82%) acquired COVID-19 from another asymptomatic person, thus making tracing difficult. We estimate an overall infection-fatality rate of 1.2% (1.0-1.3%) over the course of the outbreak, compared to a case-fatality rate of 6.3%.

### The risk of a case entering Vietnam and escaping quarantine

Available data indicate that the risk of a case entering the country is low but non-zero. Approximately 0.6% (337/55,079) of all international arrivals to airports in Vietnam tested positive to COVID-19 over April to November 2020. But data on infected passengers entering Vietnam over the period from February 1 to August 22, 2020 indicate that these measures are not failsafe. Even though 96% of infected arrivals had no known onward transmissions (Table 1), this nevertheless implies a 4% risk of an infection being released into the community despite a 14-day quarantine period, with this risk encompassing the ∼1% probability that an infected person develops symptoms only after a 14-day period,^21–23^ in addition to the probability of a failure in quarantine procedures.

**Table 1.** Statistics on the transmission dynamics of 330 COVID-19 cases arriving in Vietnam over February 1–August 22, 2020

|  |  |
| --- | --- |
| International cases with no known onward transmission | 317 (96%) |
| International cases linked to 1 known onward transmission | 10 (3%) |
| International cases linked to 2 or more known onward transmissions | 3 (1%) |

### The risk of an escaped case developing into an outbreak

If the population of Vietnam remains highly compliant with mask-wearing and other non-pharmaceutical interventions, our projections indicate that the epidemic would remain under control even if a small but steady flow of imported infections escaped quarantine into the community (Figure 3). If, on the other hand, mask usage declines as apathy increases, there is a chance that the epidemic could rebound again, with the worst-case scenarios projecting a peak of 2,500 active cases within 2 months of borders reopening (Figure 3, middle column). The worst of these outcomes could be partly mitigated if policy and/or behavior responds dynamically to news that the daily number of locally-acquired cases has exceeded a threshold, which we here assume to be 5, but in either case, the delay between when infections begin to increase and when the first cases are detected mean that significant amounts of transmission occur before policy can respond (Figure 3, final column).

**Figure 3.**
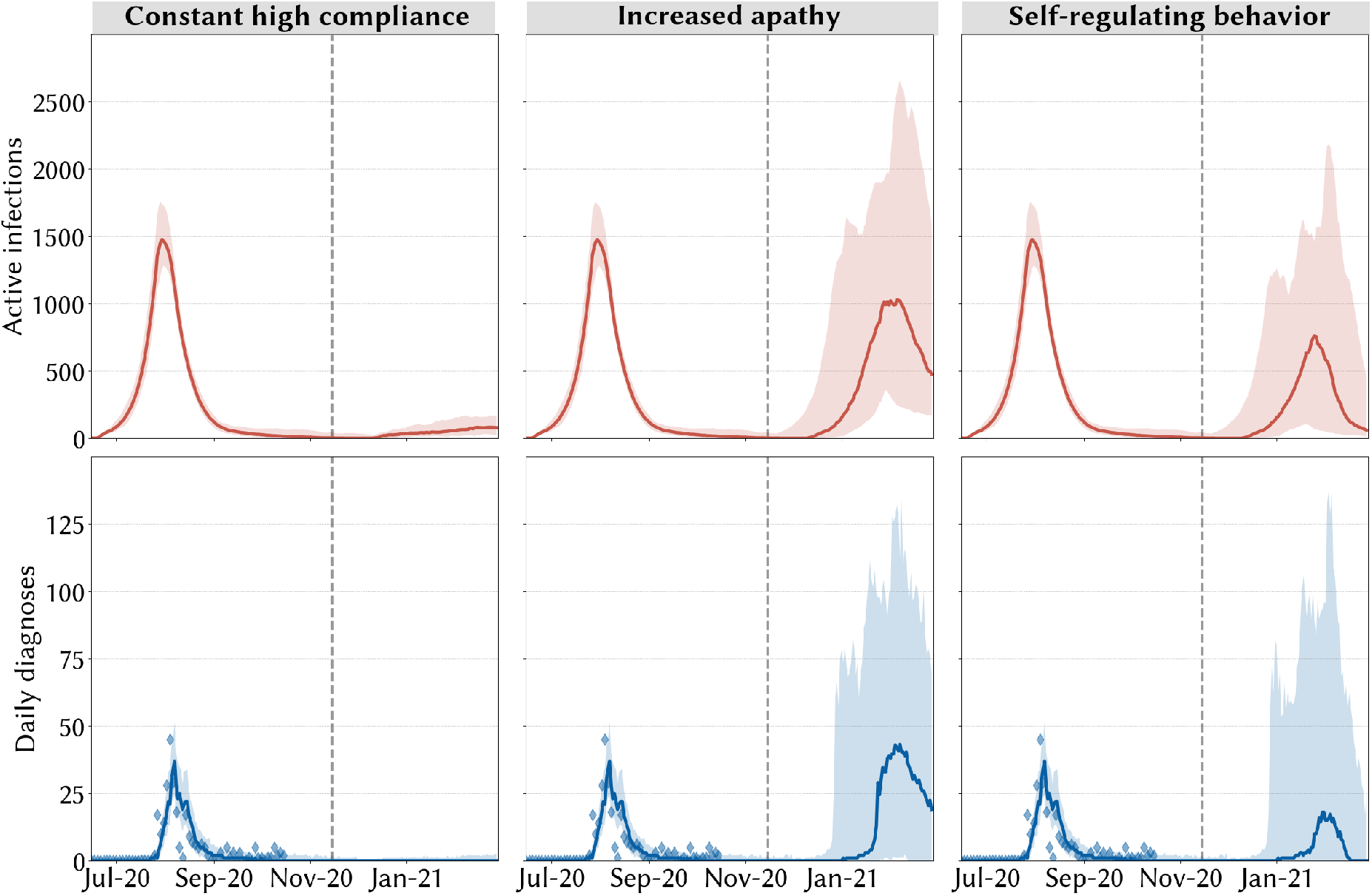
Projections of active infections and daily diagnoses under different assumptions about how people react to news of increasing case counts. In the left column, people are assumed to remain perfectly compliant with masks and other NPIs even after prolonged periods of low transmission. In the middle column, increasing apathy is assumed to lead to permanently decreased compliance with mask wearing, although government policies still mandate the closure of schools and workplaces upon the detection of new cases. In the right column, people’s compliance with mask wearing and other behavioral interventions decreases and increases as a function of reported cases.

The degree of control over future outbreaks depends to a large extent on the speed with which cases were detected, which depends on ongoing testing for those with COVID-19-like symptoms. In Figure 3, we assumed that during periods when no cases had been reported, demand for symptomatic testing would be low, with a baseline of 10% of those with symptoms seeking testing.

In Figure 4, we vary the baseline symptomatic testing rate. Across all simulations, lower testing rates would lead to a more prolonged period of increase in the epidemic before eventually reversing (Figure 4A). The difference is particularly notable as the testing rate increases from 10% to 20%, which leads to a halving of the median cumulative number of infections over the following four months from 2,110 (95% interval: 1,050-3,630) to 1,100 (570-1,670) (Figure 4B).

**Figure 4.**
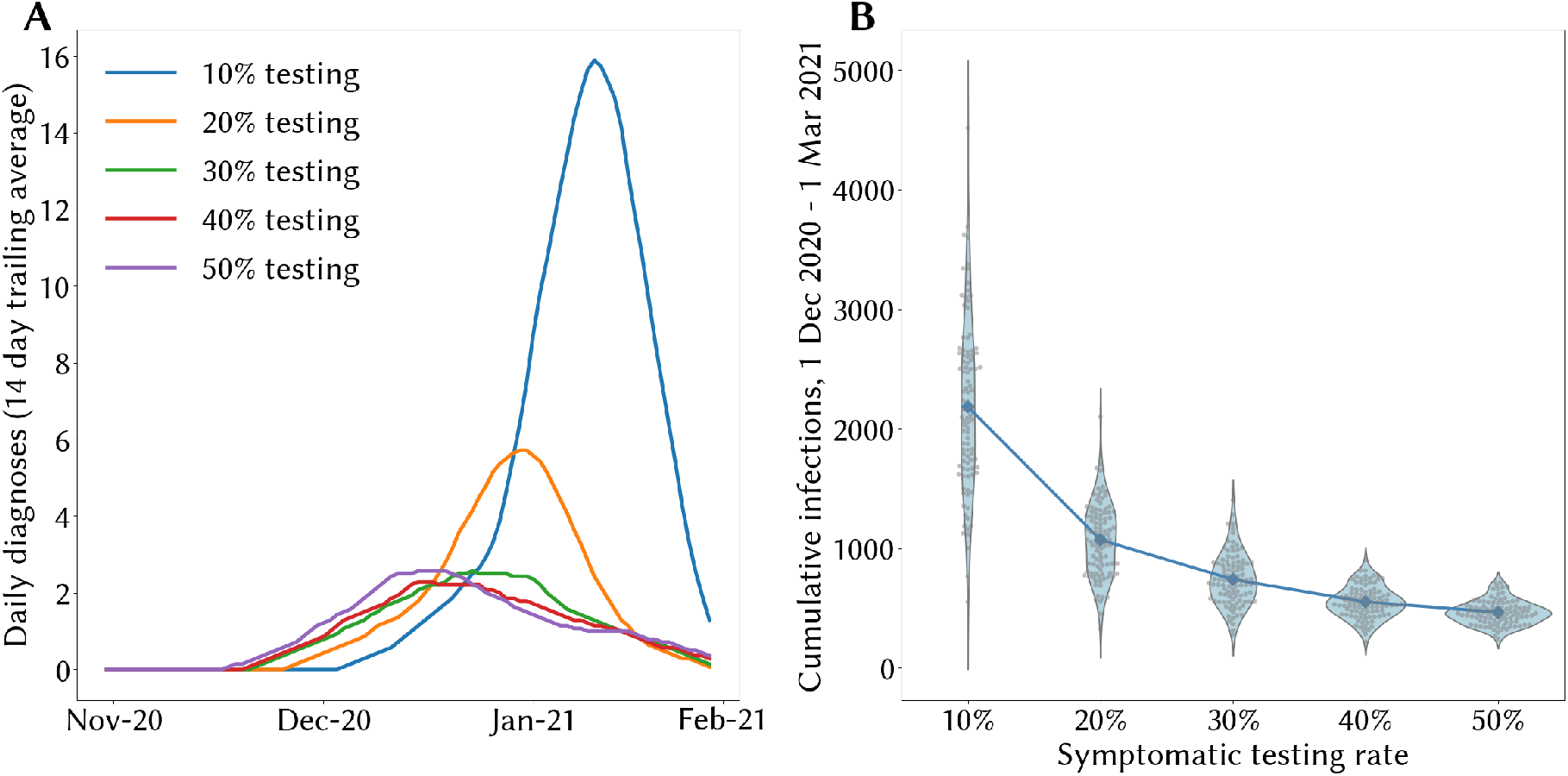
The role of routine symptomatic testing in curtailing the potential size of an outbreak in Vietnam following border reopenings. (A) median estimated trajectories of the 14-day trailing average of daily diagnoses across 100 simulations, and (B) cumulative infections over December 1, 2020 – March 1, 2021 in each simulation (grey dots, blue densities) with medians (blue dots).

## Discussion

The efficacy of Vietnam’s COVID-19 response – already well-documented in the literature^4,5^ – was demonstrated once again in the wake of the July-August outbreak in the province of Da Nang.^25^ With schools and workplaces shut down in affected areas within 3 days of cases being detected, the immediate adoption of masks, widespread testing and quarantine of potentially exposed persons, and rapid contact tracing, Vietnam was able to flatten the curve within a week of cases being detected. As remarkable as this is, however, our results suggest that there would be scope for even further improvements if those with COVID-19-like symptoms could be encouraged to seek testing even if they haven’t had a history of contact with a known case. We estimate that by the time the first cases of the Da Nang outbreak were detected, there had already been 1,480 (1,170-1,870) infections in the province. This was likely a result of a rapid influx of domestic tourists and extremely low testing rates in Da Nang. Since no quarantine or testing protocols applied to domestic travelers, these cases went undetected.

Lessons can be drawn from the experience in Da Nang as Vietnam considers reopening its borders to international travelers. Although reopening the borders will still require incoming arrivals to follow rigorous testing and quarantine protocols, our examination of incoming arrivals to Vietnam over a five month period indicated that 13/330 (4%) infected arrivals transmitted to one or more people despite these protocols. Consequently, permitting more international travelers is not a risk-free decision. Assuming that Vietnam continues to pursue a policy of COVID-19 elimination, our results indicate that any outbreaks that may result from reopening borders have a high likelihood of being contained, assuming that features of earlier responses would be repeated. However, if testing rates are low, then there would still be the potential for significant amounts of community transmission prior to the detection – and consequent containment – of new cases. We estimate that if testing rates remain at levels similar to those observed prior to the Da Nang outbreak, there could be 1,000-4,000 cumulative infections over the four months following the reopening of borders, but that a doubling of the testing rate from 10% to 20% would halve this total.

Our results on the importance of ongoing symptomatic testing even in zero- or low-transmission settings are supported by other studies in the literature. Two studies by our group examined the role of testing in very low-transmission and zero-transmission settings, respectively. The first of these found that ongoing low transmission could be largely controlled by test-and-trace strategies, but that the total number of infections over three months would be more than 100 times higher with a 50% testing rate compared to a 90% testing rate.^26^ The second estimated the probability of a single introduced case resulting in >5 cases/day within 60 days, finding a probability of ∼50% with no restrictions in place and a symptomatic testing rate of 25%, compared to 45% with a symptomatic testing rate of 50%, or 35% with a symptomatic testing rate of 75%.^27^ Elsewhere in the literature, a study by Kucharski et al found that mass random testing of 5% of the population/week combined with self-isolation, household quarantine, and manual contact tracing of all contacts would lead to a mean transmission reduction of 64%.^28^ Another study by Hellewell et al found that with 20 initial cases and 60% of contacts being traced, less than 50% of outbreaks would be controlled, even assuming that all symptomatic cases are eventually detected.^29^

Our study has a number of limitations. Firstly, since we use an agent-based model, our results are based on underlying assumptions about the ways in which these agents interact. We modelled agent interactions over four networks (households, schools, workplaces, and community), but did not explicitly model large gatherings that could potentially become super-spreader events. Such events are known to have potential for sparking outbreaks.^30,31^ Our estimates of the potential scale of an outbreak in Vietnam may therefore be conservative, especially given the proximity of the 1-week Tet holidays (Vietnamese Lunar New Year) in early February, and the National Congress of the Communist Party of Vietnam in late January 2021. Superspreading is also partly driven by overdispersion of viral load among individuals, a factor which is included in the model (e.g. in Seattle, we estimate that 50% of transmissions are caused by ∼10% of infected people^32^).

Another limitation is that we assume that the population is homogeneous in terms of behavior and quarantine compliance. In general, not including variability in model inputs means that it is also not included in the model’s outputs. For example, when models assume that mask-wearing reduces everyone’s transmission risk by a certain percentage, this population-level summary actually incorporates a range of individual behavioral changes that may adjust individual-level transmission risk by varying amounts. The possibility of pockets of variation – e.g., a single individual who happens to have a high viral load, a high number of contacts, and does not wear a mask – are significant factors in considering outbreak risk.

We further note that we do not model supply-side constraints on testing or contact tracing, but with a rapidly-growing epidemic, there is a real possibility of capacity constraints taking effect, especially for contact tracing programs, which may thus prevent tracing-based containment beyond a certain point.

Our parameter estimates for factors such as the age-dependent probability of developing symptoms or dying are based on published values that, although they represent the best information available at the time of writing,^33^ are nonetheless derived from studies that are not specific to Vietnam, and which are subject to revision as new information becomes available.

Vietnam’s success to date in pursuing a zero-COVID-19 policy is remarkable for several reasons. Compared to other countries aiming for local elimination (such as South Korea, Australia, and New Zealand), Vietnam not only has a much larger population and lower per-capita income, but it has the additional challenge of monitoring land borders. To maintain this after reopening borders to international travelers will require a continued commitment to fast and stringent policy reactions to new cases, but equally importantly, sufficient levels of symptomatic testing even among people with no known history of contact with a confirmed case. Rapid containment is only possible given the availability of real-time data on the state of the epidemic. As countries like Vietnam consider how to re-introduce international travel, the importance of routine testing as a surveillance measure will be crucial.

## Data Availability

Code for Covasim is available from https://github.com/institutefordiseasemodeling/covasim. The code and data used to run all the analyses that are included in this article are available from the corresponding author upon request during the article review period, and will be placed in a public GitHub repository after manuscript acceptance.

## Supplementary materials

- Table S1: parameters and intervention effects used in the model for Vietnam

## Acknowledgements

The authors would like to thank additional members of the Institute for Disease Modeling team who contributed to the base Covasim model. We also thank frontline colleagues from parts of Vietnam who have devoted their lives to taking care of COVID-19 patients and their enormous efforts to trace, quarantine, and test countless close contacts of cases and at-risk individuals over the course of the present COVID-19 pandemic. Their clinical and epidemiological data collection and data sharing were essential for this modelling exercise. The authors appreciate Dr Tu N Le and Dr Thinh V Nguyen at the Pasteur Institute of Ho Chi Minh City for management of model data inputs.

## Supplementary materials

**Table S1:**
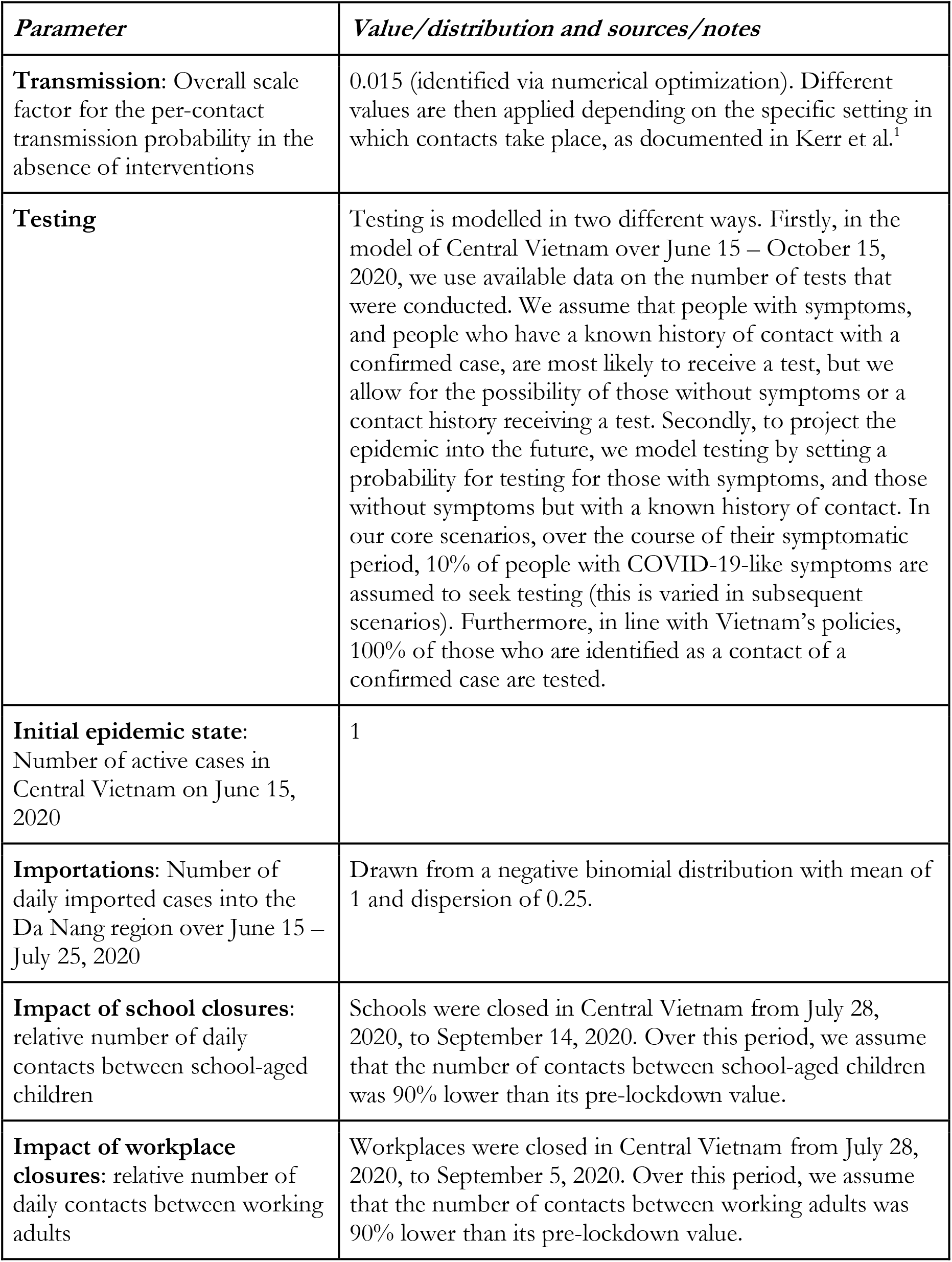
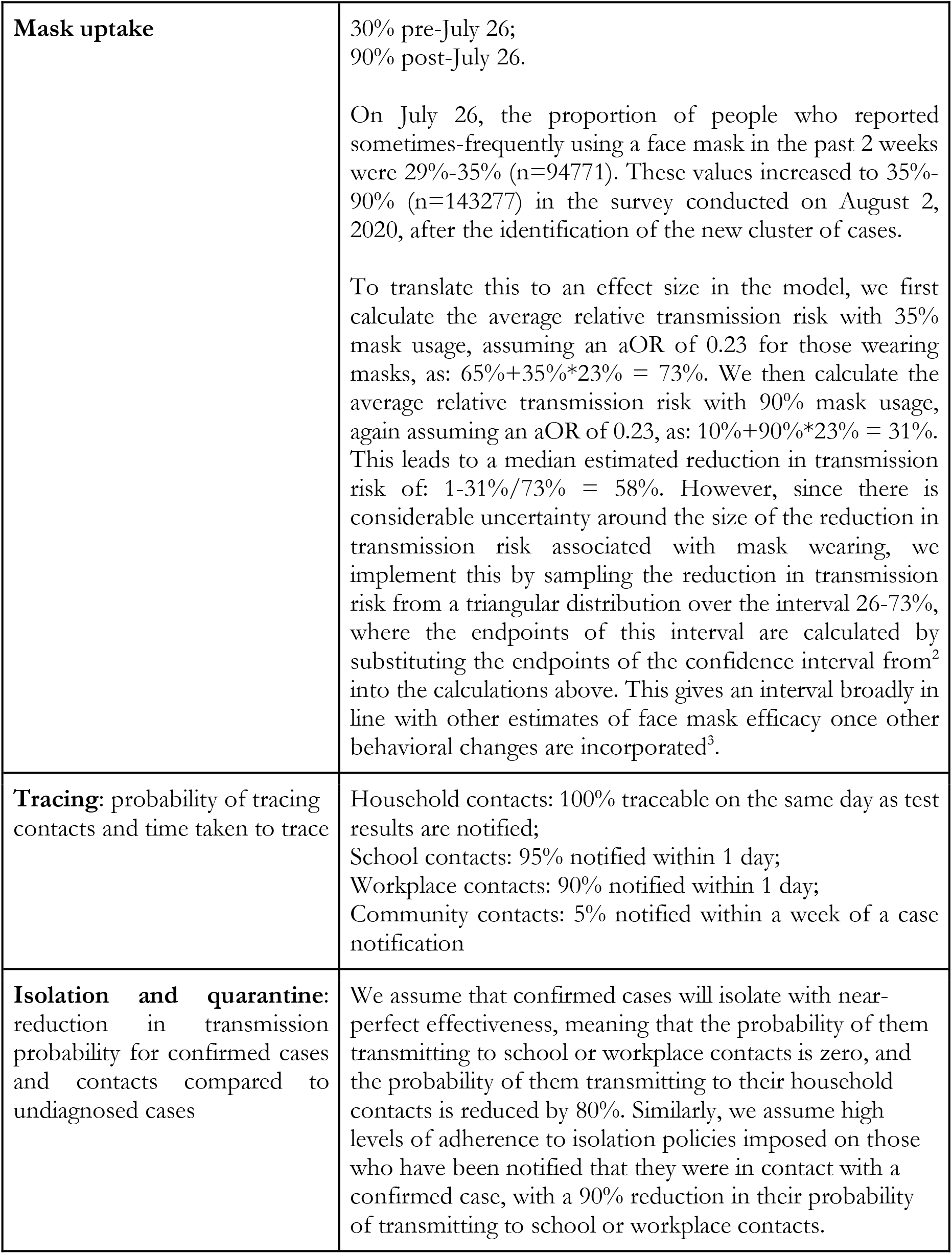
list of parameters used in the calibrated Covasim model for Central Vietnam. For other model parameters, we use the default Covasim values^1^.

